# Total Gleason Pattern 4 Length Outperforms Grade Group and Clinical Models in Predicting Oncologic Outcomes in Grade Group 2-4 Prostate Cancer

**DOI:** 10.64898/2026.02.27.26347289

**Authors:** Nicholas A. Pickersgill, Sean A. Fletcher, Nate Aiken, Melissa J. Assel, Nicole Liso, Victor E. Reuter, Andrew J. Vickers, Behfar Ehdaie, Samson W. Fine

## Abstract

**Background and Objective:** Risk stratification in localized prostate cancer relies primarily on Grade group (GG). In GG2-4 disease, risk assignment depends on the proportions of pattern 3 and pattern 4. We hypothesized that total pattern 4 length on biopsy would better predict oncologic outcome than GG, percent pattern 4, and multivariable models (“nomograms”) based on clinical variables.

**Methods:** We identified 2499 patients with GG2-4 prostate cancer on biopsy who underwent radical prostatectomy. Discrimination for predictors was calculated for adverse pathologic stage (seminal vesicle invasion or lymph node invasion) and biochemical recurrence (BCR).

**Key Findings and Limitations:** Total pattern 4 length for the case demonstrated the highest discrimination for adverse pathologic stage in comparison with GG (AUC 0.779 vs 0.658; p<0.0001), percent pattern 4 (0.719), and a model including prostate-specific antigen level, clinical stage, GG, PI-RADS score, and number of positive cores (0.762). Results were similar for BCR, with total pattern 4 length outperforming GG (C-index 0.716 vs 0.662), percent pattern 4 (0.695), and the clinical model (0.699). Neither mm of pattern 3 nor the clinical model added discrimination to total mm of pattern 4.

**Conclusions and Clinical Implications:** Total length of Gleason pattern 4 on biopsy best predicts oncologic outcome in GG2-4 prostate cancer. Other common clinicopathologic variables do not further aid discrimination. Further research is warranted to determine the optimal method for quantifying pattern 4 before incorporation into risk stratification algorithms.

- **What does the study add?**: Patients with Grade group 2-4 prostate cancer constitute both the largest group and the one in which treatment decision-making is most difficult. For such patients, total length of Gleason pattern 4 on biopsy predicted oncologic outcomes better than Grade group or multivariable models including the standard predictors of stage, grade, PSA, PI-RADS and number of positive cores. Neither mm of pattern 3 nor the standard predictors add discrimination once total length of pattern 4 is known.
- **Patient Summary**: Treatment decisions in prostate cancer are often determined by the ratio of pattern 4 to pattern 3 disease. We showed that, in GG2-4 disease, using the total amount of pattern 4 for the case better predicts risk and therefore provides a better basis for treatment decisions.

**Take Home Message:** In Grade group 2-4 prostate cancer, total Gleason pattern 4 length for the case is a stronger predictor of adverse pathologic stage and biochemical recurrence than Grade group and other standard clinical variables. Further research is warranted to determine the optimal method for quantifying pattern 4 before incorporation into risk stratification algorithms.

## Introduction

Risk stratification and clinical decision-making in localized prostate cancer are largely determined by Grade group (GG). In men with GG2-4 disease, which constitutes the group that is both the largest and the one in which treatment decision-making is most difficult, GG relies upon the relative proportion of more aggressive (Gleason pattern 4) to indolent (Gleason pattern 3) disease^1,2^. Use of proportions is distinctively uncommon in cancer pathology and can lead to counterintuitive results. For instance, a patient with a very large amount of pattern 4, but slightly more pattern 3, is paradoxically classified as lower risk (GG2) than an individual with a small focus of pattern 4 but no pattern 3 (GG4).

Given that pattern 3 is biologically indolent^3^, it remains unclear as to why it should play a signficant role in determining prognosis and treatment. Several studies have provided preliminary data supporting such an approach, with quantification of pattern 4 (whether mm on biopsy or cc on prostatectomy specimen) providing superior risk discrimination to Gleason score for predicting adverse pathology, biochemical recurrence (BCR), and even metastatic progression^4–7^. Although promising, this literature has been limited by focus on GG2 disease only^4,5^, quantification of radical prostatectomy pattern 4 volume^6^ (which holds less value for treatment decision-making), or relatively limited sample size (n=446)^7^.

In 2015, pathologists at Memorial Sloan Kettering Cancer Center (MSKCC) began to routinely report percentage of pattern 4 for all GG2 cores. This was later expanded to include GG3 cores as well. As each cancer-bearing core also had length of cancer in mm documented, it allows us to calculate total mm of pattern 4 for each case among a large group of contemporary patients with GG2-4 disease. The large sample size allows us to ask not only whether quantitative pattern 4 length offers superior predictive discrimination to GG, but also how it compares to multivariable models (“nomograms”) including standard clinical variables. In particular, we were interested in determining whether additional information on mm of pattern 3 or routine contemporary clinical variables add prognostic value once total length of pattern 4 is known.

## Methods

We identified patients with clinically localized or locoregional prostate cancer that underwent a radical prostatectomy (RP) and pelvic lymph node dissection at our institution between 2013 and 2024. We had different windows of inclusion (years) depending on the GG. Prostate biopsy cores with GG2 had percentage of pattern 4 documented from mid-2015 onward and GG3 from 2018 onward. Patients with GG4 only on biopsy between 2013 and 2018 were also included. We excluded patients with any pattern 5. All biopsy pathology was reviewed by dedicated genitourinary pathologists at our institution.

To calculate mm of pattern 4 on biopsy, we used routinely collected data from the pathology report on percentage of pattern 4 and mm of cancer in each cancer-bearing core, multiplying the two to obtain mm of pattern 4. Total mm of pattern 4 was calculated as the sum of the lengths of pattern 4 on systematic biopsy plus the mean length of the cores from each MRI target. For instance, if there were two positive cores from systematic biopsy each measuring 1 mm in length, and the three cores taken from the same MRI target had 0 mm, 1 mm and 2 mm, total mm of pattern 4 would be calculated as 2 mm (the sum of the systematic cores) plus 1 mm (the mean of the targeted cores) to give a total of 3 mm. To calculate mm of pattern 3, we subtracted mm of pattern 4 from total mm of cancer. As one comparator to total mm of pattern 4, we also calculated the mean percent pattern 4 in cores with any pattern 4 in systematic and MRI-targeted biopsies separately and used the maximum mean percentage for further analysis.

Adverse pathologic stage was defined as either seminal vesicle invasion (SVI) or lymph node involvement (LNI). Extraprostatic extension (EPE) was added to this definition in a sensitivity analysis. Biochemical recurrence (BCR) was defined as a prostate-specific antigen (PSA) level greater than 0.2 ng/mL with a second confirmatory rise.

The discrimination of total mm of pattern 4 for the case was compared with GG, maximum percentage pattern 4 in any one core, and a multivariable model of standard clinical predictors: clinical stage (T1, T2a, T2b, T2c, T3/4), GG, PSA, PI-RADS score^8^ (1/2, 3, 4, 5) and number of positive systematic biopsy cores. Predictors of adverse pathologic stage were evaluated by logistic regression, with discrimination estimated by the area-under-the-curve (AUC), corrected for overfit by 10-fold cross-validation for multivariable models. Differences in discrimination were by Delong, Delong and Clarke-Pearrson^9^. A comparable approach was taken for BCR using Cox regression and the concordance index (C-Index), with p-values for difference in C-Index between models calculated by bootstrapping to determine the standard error for the difference using a t-test.

All analyses were conducted using R version 4.4.0 with the gtsummary (v1.7.2) and tidyverse (v2.0.0) packages.

## Results

We excluded 164 patients who had inadequate biopsy sampling (<10 cores), 46 who underwent saturation biopsy (>20 systematic cores), and 33 with evidence of pattern 5. An additional 481 patients were excluded due to insufficient documentation to determine core location or percent pattern 4. After applying all exclusion criteria, the final analytic cohort comprised 2499 patients. Baseline patient and disease characteristics are summarized in Table 1.

**Table 1.**
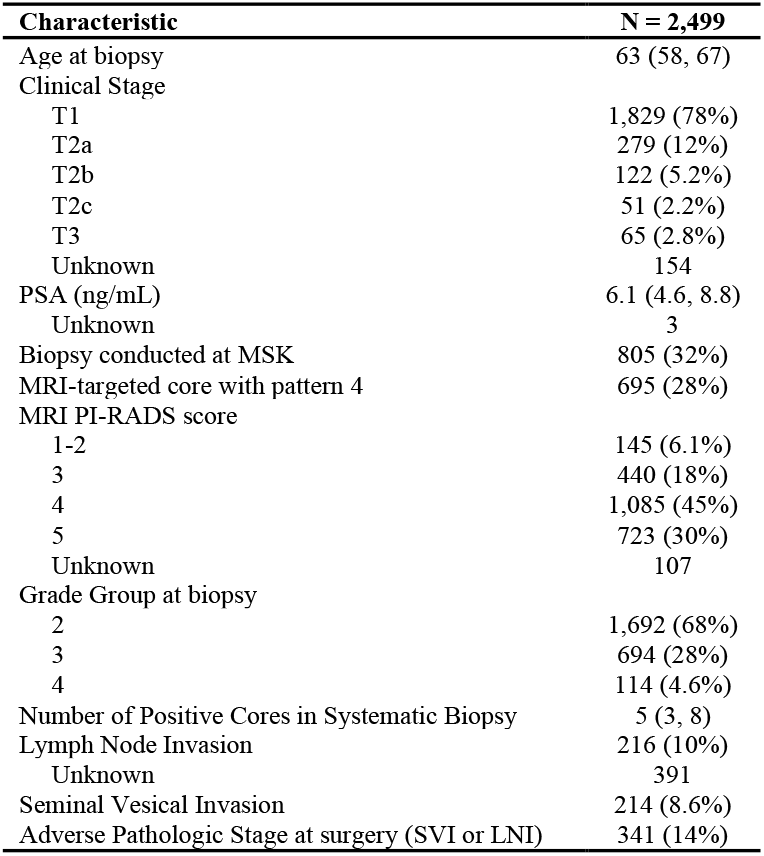
Patient and disease characteristics. Results are presented as median (interquartile range) and frequency (%). PSA = prostate-specific antigen; MRI = magnetic resonance imaging.

| Characteristic | N = 2,499 |
| --- | --- |
| Age at biopsy | 63 (58, 67) |
| Clinical Stage |  |
| T1 | 1,829 (78%) |
| T2a | 279 (12%) |
| T2b | 122 (5.2%) |
| T2c | 51 (2.2%) |
| T3 | 65 (2.8%) |
| Unknown | 154 |
| PSA (ng/mL) | 6.1 (4.6, 8.8) |
| Unknown | 3 |
| Biopsy conducted at MSK | 805 (32%) |
| MRI-targeted core with pattern 4 | 695 (28%) |
| MRI PI-RADS score |  |
| 1-2 | 145 (6.1%) |
| 3 | 440 (18%) |
| 4 | 1,085 (45%) |
| 5 | 723 (30%) |
| Unknown | 107 |
| Grade Group at biopsy |  |
| 2 | 1,692 (68%) |
| 3 | 694 (28%) |
| 4 | 114 (4.6%) |
| Number of Positive Cores in Systematic Biopsy | 5 (3, 8) |
| Lymph Node Invasion | 216 (10%) |
| Unknown | 391 |
| Seminal Vesical Invasion | 214 (8.6%) |
| Adverse Pathologic Stage at surgery (SVI or LNI) | 341 (14%) |

Results for adverse pathologic stage are shown in Table 2. Total length of pattern 4 in mm for the case demonstrated substantially better discrimination for adverse pathologic stage than GG alone (AUC 0.783 vs 0.634; p<0.001), and was also superior to the maximum percent pattern 4 in either systematic or targeted cores (AUC improvement 0.060, p<0.001). Total millimeters of pattern 4 also had higher discrimination than a model with clinical predictors, although this difference was not statistically significant (AUC difference 0.025, p=0.2).

**Table 2.** Discrimination of Gleason pattern 4 length versus other predictors. Values are presented as area under the curve or C-index (95% confidence interval). Multivariable models corrected for optimism using 10-fold cross validation. Standard clinical predictors were clinical stage (T1, T2a, T2b, T2c, T3/4), Grade group, PSA, PI-RADS score (1/2, 3, 4, 5) and number of positive systematic biopsy cores. PLND = pelvic lymph node dissection; SVI = seminal vesicle invasion; LNI = lymph node invasion; EPE = extraprostatic extension.

| Endpoint | Patient Group | Total mm Pattern 4 | Grade Group | Max % Pattern 4 | Standard Clinical Predictors | Standard Clinical Predictors plus mm Pattern 4 |
| --- | --- | --- | --- | --- | --- | --- |
| Primary analysis: SVI or LNI | All | 0.783 (0.754, 0.812) | 0.634 (0.604, 0.664) | 0.707 (0.679, 0.736) | 0.758 (0.727, 0.789) | 0.777 (0.746, 0.809) |
| Biochemical recurrence | All | 0.707 (0.679, 0.735) | 0.636 (0.613, 0.659) | 0.680 (0.652, 0.708) | 0.685 (0.657, 0.713) | 0.677 (0.649, 0.705) |
| Sensitivity analyses |  |  |  |  |  |  |
| SVI or LNI or EPE | All | 0.706 (0.684, 0.727) | 0.573 (0.554, 0.592) | 0.629 (0.606, 0.652) | 0.729 (0.708, 0.749) | 0.747 (0.726, 0.767) |
| SVI or LNI | PLND | 0.788 (0.759, 0.818) | 0.643 (0.611, 0.675) | 0.716 (0.688, 0.745) | 0.765 (0.733, 0.797) | 0.782 (0.75, 0.814) |
| SVI or LNI or EPE | PLND | 0.710 (0.687, 0.732) | 0.580 (0.559, 0.601) | 0.636 (0.612, 0.661) | 0.733 (0.710, 0.755) | 0.748 (0.727, 0.77) |

Table 3 shows the multivariable model to predict advanced stage using total mm of pattern 4 and clinical predictors. Although some of the clinical predictors were statistically significant, they did not add to the discrimination of total length of pattern 4 alone (Table 2). These findings were broadly consistent in sensitivity analyses that included EPE as part of the adverse pathologic stage endpoint or excluded patients who did not undergo pelvic lymph node dissection, although the contribution of clinical predictors was modestly greater when the less aggressive endpoint of EPE was included.

**Table 3.**
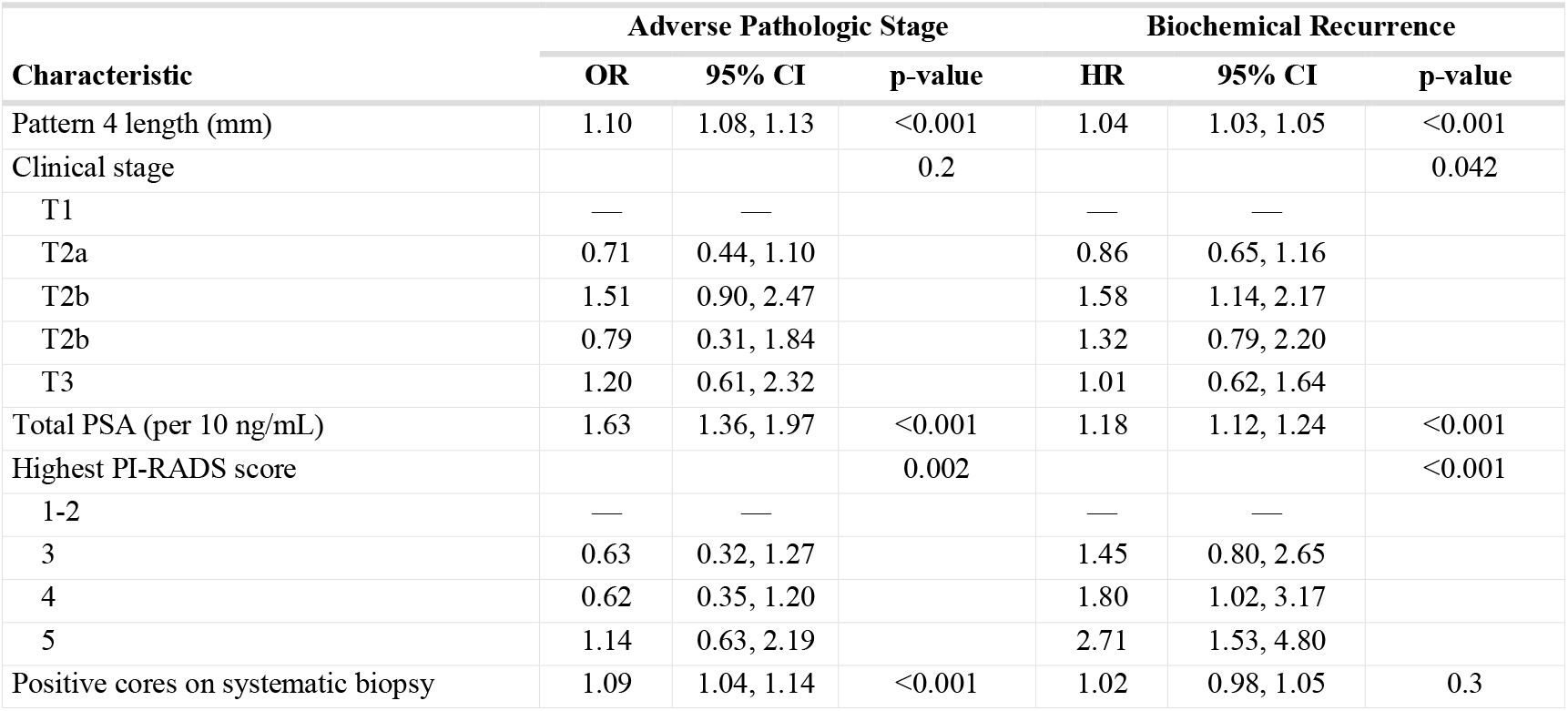
Individual multivariable models of clinical predictors plus length of pattern 4 in mm. PSA = prostate-specific antigen; PI-RADS = Prostate Imaging Reporting and Data System; CI = confidence interval; OR = odds ratio; HR = hazard ratio.

| Characteristic | Adverse Pathologic Stage |  |  | Biochemical Recurrence |  |  |
| --- | --- | --- | --- | --- | --- | --- |
|  | OR | 95% CI | p-value | HR | 95% CI | p-value |
| Pattern 4 length (mm) | 1.10 | 1.08, 1.13 | <0.001 | 1.04 | 1.03, 1.05 | <0.001 |
| Clinical stage |  |  | 0.2 |  |  | 0.042 |
| T1 | — | — |  | — | — |  |
| T2a | 0.71 | 0.44, 1.10 |  | 0.86 | 0.65, 1.16 |  |
| T2b | 1.51 | 0.90, 2.47 |  | 1.58 | 1.14, 2.17 |  |
| T2b | 0.79 | 0.31, 1.84 |  | 1.32 | 0.79, 2.20 |  |
| T3 | 1.20 | 0.61, 2.32 |  | 1.01 | 0.62, 1.64 |  |
| Total PSA (per 10 ng/mL) | 1.63 | 1.36, 1.97 | <0.001 | 1.18 | 1.12, 1.24 | <0.001 |
| Highest PI-RADS score |  |  | 0.002 |  |  | <0.001 |
| 1-2 | — | — |  | — | — |  |
| 3 | 0.63 | 0.32, 1.27 |  | 1.45 | 0.80, 2.65 |  |
| 4 | 0.62 | 0.35, 1.20 |  | 1.80 | 1.02, 3.17 |  |
| 5 | 1.14 | 0.63, 2.19 |  | 2.71 | 1.53, 4.80 |  |
| Positive cores on systematic biopsy | 1.09 | 1.04, 1.14 | <0.001 | 1.02 | 0.98, 1.05 | 0.3 |

A total of 517 patients experienced BCR; median follow-up for patients without BCR was 2.3 years. Results for BCR mirrored those observed for adverse pathologic stage. Total pattern 4 length demonstrated higher discrimination than GG (C-index 0.707 vs 0.636, p<0.001) and maximum percent pattern 4 in either the systematic or targeted biopsy (0.680, p=0.003), and also outperformed the clinical model (0.696, p=0.2). As with adverse pathologic stage, while some clinical variables were statistically significant in a multivariable model including total pattern 4 length (Table 3), their inclusion did not improve model discrimination (Table 2).

To explore this result further, we explored the association between total length of pattern 4 and each of PSA and PI-RADS, the only two clinical predictors with consistent, statistically significant associations with outcome in the multivariable models. We found total mm of pattern 4 in patients with a PSA ≥10 ng/mL was approximately twice that of patients with PSA <10 ng/mL (7 mm vs 3.6 mm); for PI-RADS, mean total mm of pattern 4 increased from 1.8 for PI-RADS ≤3 to 3.1 for PI-RADS 4 and to 7.7 for PI-RADS 5. The correlation between total mm pattern 4 and these clinical predictors helps explain why they did not add information beyond total mm pattern 4 alone.

We also evaluated whether incorporating mm of pattern 3 improved prediction beyond total mm of pattern 4. Although pattern 3 length was statistically significant in the model predicting adverse pathologic stage (p<0.001), it did not improve discrimination compared with total pattern 4 length alone (AUC 0.770 for mm pattern 4 plus mm pattern 3 vs 0.783 for mm pattern 4 alone). For BCR, pattern 3 length was not statistically significant (p=0.10). Results were unchanged in a sensitivity analysis which calculated mm of pattern 3 only in cores in which there was pattern 4, and excluding those cores with only pattern 3.

## Discussion

In this large contemporary cohort of men with GG2-4 prostate cancer undergoing radical prostatectomy, we found that total length of pattern 4 on biopsy provided substantially better discrimination for adverse pathologic stage and BCR than GG, maximum percent pattern 4 on any one core, or comprehensive clinical models incorporating PSA, clinical stage, GG, PI-RADS score, and biopsy core metrics. Importantly, once total pattern 4 length was known, no additional variable meaningfully improved discrimination.

These findings add to a growing body of evidence demonstrating the superiority of absolute pattern 4 burden over GG and percentage pattern 4 for prognostication in GG 2-4. Early work from our group showed that total mm of pattern 4 across all biopsy cores provided greater net benefit for predicting adverse pathology than GG or percent pattern 4 in GG 2 disease.^4^ A subsequent study of a cohort with GG 2 disease demonstrated that increasing pattern 4 length was strongly associated with BCR following RP, with a marked increase in recurrence risk per unit increase in pattern 4 burden.^5^ More recently, Scuderi et al. evaluated RP specimens and showed that absolute pattern 4 volume was more strongly associated with advanced pathologic stage and BCR than GG or percent pattern 4, and that pattern 3 volume did not add meaningful prognostic value once pattern 4 was accounted for.^6^ Olivier et al. extended these observations to MRI-era diagnostics, demonstrating total pattern 4 length on biopsy and pattern 4 volume derived from combining percent pattern 4 with MRI lesion size outperformed GG and widely used clinical risk scores in predicting metastatic recurrence (C-index of 0.77-0.80 for pattern 4 quantification vs 0.69 for Gleason score and 0.72 for CAPRA)^7^. Across these cohorts and endpoints, there is a consistent pattern: risk increases incrementally with the amount of pattern 4 present in a case, whereas amount of pattern 3 contributes little independent prognostic information once pattern 4 has been quantified.

Our results replicate these findings and add several important analyses. In particular, not only do we show that total pattern 4 length outperforms a comprehensive contemporary clinical model including PI-RADS, which has been increasingly incorporated into risk stratification models, but also the variables included in these models do not improve discrimination once total mm of pattern 4 is known. In other words, there is no prognostic information captured in variables such as PSA, clinical stage, number of cores, and PI-RADS that is not already captured in total mm of pattern 4.

There are sound biological explanations for our findings. With respect to the superiority of pattern 4 quantification to either GG or percent pattern 4, the latter both depend on the amount of pattern 3 present. As pattern 3 is biologically indolent and incapable of metastatic spread,^3,10,11^ it is perhaps intuitive that risk should not depend on the amount of pattern 3. Moreover, the relationship between amount of pattern 3 and risk inherent in the Gleason score/GG is opposite to that expected. For two patients with the same amount of pattern 4 in a given core, the patient with more pattern 3, and therefore more cancer overall, will be assigned a lower risk. Indeed, our finding that pattern 3 length does not aid prediction once pattern 4 length is known in GG2-4 disease strongly indicates that at the case level, it is the amount of high-grade disease and not the Gleason score/GG that best predicts outcomes. With respect to the superiority of pattern 4 quantification to standard predictors – and the failure of those predictors to improve prediction once total pattern 4 is known – a biological explanation is straightforward: PSA levels are largely driven by pattern 4 burden^12^, while clinical stage, number of positive cores, and PI-RADS primarily reflect overall tumor volume rather than the amount of aggressive histology.

Linear length of pattern 4 (at the core or case level) is a calculated value, and is not routinely stated in pathology reports. For cores with GG2 and GG3, pathologists will measure the cancer in mm and provide a percentage pattern 4. Length of pattern 4 for the core can be obtained by multiplying those two measures. For cores with GG4, the entire length of cancer provided is presumed to be pattern 4. The strength of our findings regarding total length of pattern 4 suggest strongly that pathologists report mm of cancer for each cancer-bearing core, to facilitate utilization of calculated parameters, such as total length pattern 4, for risk stratification.

Likewise, further research on pattern 4 quantification is required before incorporation into risk stratification and to guide treatment decision making. There is considerable variation in the methods used by pathologists to quantify pattern 4, with up to four-fold variation in measures of pattern 4 between pathologists evaluating the same specimen^13^. More specifically, how pathologists consider inter-focal stroma in tumor measurement and consequently, in pattern 4 quantification, is variable^14,15^.

The use of MRI-targeted biopsy introduces additional complexity. Targeted biopsy preferentially samples regions enriched for higher-grade disease and may overestimate gland-wide pattern 4 burden, whereas systematic biopsy may miss pattern 4 foci outside MRI-visible lesions.^7,16–19^ In this study, our approach was to sum pattern 4 length across systematic cores and average across targeted cores, but it is not known whether this is the optimal strategy. How best to integrate systematic and targeted sampling, particularly in multifocal or heterogeneous disease, remains an open methodological question and an area for future investigation. Similarly, incorporation of architectural subtypes of pattern 4, such as cribriform morphology^20–24^ into pattern 4 assessment and risk stratification is not standard. Ultimately, how to formally translate pattern 4 quantification into clinical decision-making has yet to be established. For instance, existing guidelines match GG to duration of androgen deprivation therapy in patients undergoing radiotherapy; such guidelines have yet to be modified for parameters such as total length of pattern 4.

In conclusion, in Grade group 2-4 prostate cancer, risk of adverse pathologic stage and biochemical recurrence is driven primarily by the absolute burden of Gleason pattern 4 disease. Total length of pattern 4 alone predicts outcome better than Grade group or multivariable models incoroporating common clinical, pathologic, and radiologic variables and discrimination is not enhanced by adding other predictors. Further research is warranted to determine the optimal method for quantifying pattern 4 before incorporation into risk stratification algorithms.

## Data Availability

All data produced in the present study are available upon reasonable request to the authors.

